# Knowledge, Practice and associated factors towards the Prevention of COVID-19 among high-risk groups: A cross-sectional study in Addis Ababa, Ethiopia

**DOI:** 10.1101/2020.08.14.20172429

**Authors:** Atkure Defar, Gebeyaw Molla, Saro Abdella, Masresha Tessema, Muhammed Ahmed, Ashenif Tadele, Fikresilassie Getachew, Bezawit Hailegiorgis, Eyasu Tigabu, Sabit Ababor, Ketema Bizuwork, Assefa Deressa, Geremew Tassaw, Addisu Kebede, Daniel melese, Andargachew Gashu, Kirubel Eshetu, Adamu Tayachew, Mesfin Wossen, Abduilhafiz Hassen, Shambel Habebe, Zewdu Assefa, Aschalew Abayneh, Ebba Abate, Getachew Tollera

**Affiliations:** Ethiopian Public Health Institute, Addis Ababa, Ethiopia; Department of Paediatrics and Child Health, School of Medicine, College of Medicine and Health Sciences, University of Gondar, Gondar, Ethiopia; School of Nursing and Midwifery College of Health Sciences, Addis Ababa University, Addis Ababa, Ethiopia

**Keywords:** COVID-19, Ethiopia, Knowledge, Practice, Prevention

## Abstract

**Background:** Coronavirus disease (COVID-19) is a highly transmittable virus that continues to disrupt livelihoods, particularly those of low income segments of society, around the world. In Ethiopia, more specifically in the capital city of Addis Ababa, a sudden increase in the number of confirmed positive cases in high-risk groups of the community has been observed over the last few weeks of the first case. Therefore, this study aims to assess knowledge, practices and associated factors that can contribute to the prevention of COVID-19 among high-risk groups in Addis Ababa.

**Methods:** A cross-sectional in person survey (n=6007) was conducted from 14-30 April, 2020 following a prioritization within high-risk groups in Addis Ababa. The study area targeted bus stations, public transport drivers, air transport infrastructure, health facilities, public and private pharmacies, hotels, government-owned and private banks, telecom centers, trade centers, orphanages, elderly centers, prison, prisons and selected slum areas where the people live in a crowed. A questionnaire comprised of four sections (demographics, knowledge, practice and reported symptoms) was used for data collection. The outcomes (knowledge on the transmission and prevention of COVID-19 and practices) were measured using four items. A multi variable logistic regression was applied with adjustment for potential confounding.

**Results:** About half (48%, 95% CI: 46-49) of the study participants had poor knowledge on the transmission mode of COVID-19 whereas six out of ten (60%, 95% CI: 58-61) had good knowledge on prevention methods for COVID-19. The practice of preventive measures towards COVID-19 was found to be low (49%, 95% CI: 48-50). Factors that influence knowledge on COVID-19 transmission mechanisms were female gender, older age, occupation (health care and grocery worker), lower income and the use of the 8335 free call centre. Older age, occupation (being a health worker), middle income, experience of respiratory illness and religion were significantly associated with being knowledgeable about the prevention methods for COVID-19. The study found that occupation, religion, income, knowledge on the transmission and prevention of COVID-19 were associated with the practice of precautionary measures towards COVID-19.

**Conclusion:** The study highlighted that there was moderate knowledge about transmission modes and prevention mechanism. Similarly, there was moderate practice of measures that contribute towards the prevention of COVID-19 among these priority high risk communities of Addis Ababa. There is an urgent need to fill the knowledge gap in terms of transmission mode and prevention methods of COVID-19 to improve preventions practices and control the spread of COVID-19. Use of female public figures and religious leaders could support the effort towards the increase in awareness.

## Background

Coronaviruses are a large family group of viruses that cause illnesses that range from the common cold to more severe diseases found in both animals and humans (1). A sudden outbreak of coronavirus disease 2019 (COVID-19) caused by infection with severe acute respiratory syndrome coronavirus 2 (SARS-CoV-2) began in December 2019 in Wuhan City, Hubei Province, China (2). The most recent novel coronavirus, currently known as COVID-19 virus, had not been detected before and could be a new strain of coronavirus that has no antecedent known in humans (3).

Fever, dry cough, fatigue, and multiple systemic illnesses including respiratory (shortness of breath, sore throat, rhinorrhea, hemoptysis, and chest pain), gastrointestinal (diarrhoea, nausea, and vomiting), musculoskeletal (muscle ache), and neurologic (headache or confusion) were the symptoms reported in the first cases in China (1,4). The best estimate of the incubation period of the disease ranges from 1 to 14 days (3), it causes a huge burden on the healthcare system and is a global public health emergency for all countries in the world (3,5).

According to the Worldometer (As of August 4, 2020, 7 pm), COVID-19 has spread to more than 210 countries and territories and accounted for 18,543,936 confirmed cases. Of these, 63.43% (11,762,600) cases recovered and 3.77% (699,987) of total reported cases have died (6). In Africa, according to the Center for Disease Control (CDC) dashboard, since the first case was registered in 14 February 2020 in Egypt, 968,020 cases, 629, 612 recoveries and 20, 612 deaths have been reported. According to the Ministry of Health and the Ethiopian Public Health Institute’s COVID-19 pandemic preparedness and response daily situation report (4 August 2020), since the first COVID-19 case was reported on 13 March, 2020, a total of 19, 877 COVID-19 confirmed positive cases have been registered; out of these, 41.45%(8249) have recovered and 1.72%(343) have died (7).

The global community is struggling to slow down and eventually halt the spread of COVID-19 that has claimed thousands of lives and sickened tens of thousands through improving the knowledge and practice of COVID-19 prevention methods, testing and screening (8,9).

WHO reports that the best way to prevent and slow down the transmission of COVID-19 is to accurately and widely inform the public about the disease, the causes, mode of transmission, and simple prevention methods such as handwashing with soap or use of hand sanitizers, maintaining social distance and staying home to remain protected from the infection (10). On the other hand, poor hand hygiene practices, overcrowding and close physical contacts like handshaking contribute to the fast spread of the virus within a very short period (11). Implementing personal hygiene and public health interventions especially in priority high-risk groups is necessary to curb the spread of coronavirus (12). Therefore, enhancing the community’s knowledge and practice of COVID-19 symptoms & prevention methods will have a significant contribution to reduce the spread of the outbreak. A study conducted in United States of America on awareness, attitudes, and actions related to COVID-19 revealed that seven out of ten participants (71.7%) correctly identified three COVID-19 symptoms and 69.8% were able to identify three prevention methods (13). Studies in China and the United Arab Emirates revealed that 73.8% of health workers identified fever, cough, sore throats and shortness of breath as symptoms of COVID-19 (14,15), while, 98% of health workers suggested washing hands with soap and water, social distance and using face masks could help in the prevention of disease transmission (14). In line with this, 96% of Chinese residents had mentioned fever, fatigue, dry cough, and myalgia as the main clinical symptoms of COVID-19 (16). Another study in Bangladesh also indicated that 98.7% of students believed that handwashing with soap and water followed by avoiding touching your nose, mouth and eyes with unwashed hands, the use of face mask/tissue when coughing or sneezing and wearing a clean surgical mask during their respiratory illness as the prevention methods for COVID-19 (17).

Ethiopia is currently showing a high commitment to prevent and slow down the COVID-19 pandemic before it causes significant health damages and an economic crisis in the community. Case identification, contact tracing, isolation and mandatory quarantine, large scale screening in high-risk groups are the actions being taken to control the spread of the disease (11). Controlling the spread of infection in high-risk groups must be the target in the containment strategy of COVID-19 responses. To our knowledge, no study has assessed the knowledge and practices towards the prevention of COVID-19 among high-risk groups. Therefore, this study aimed to assess the knowledge, practices, and associated factors towards the prevention of COVID-19 among the high-risk groups of Addis Ababa community.

## Methods

### Study area and period

The study was conducted in Addis Ababa, the capital city of Ethiopia. Addis Ababa is often referred to as “the political capital of Africa” for its historical, diplomatic and political significance for the continent. The largest city in the country, with a total population of 4,793,699 (52% Female Vs 48% Male). The religious composition of the population showed the overwhelming majority to be Orthodox Christian (81.8%), followed by Muslim (12.7%), Protestant (3.9%) and Catholic (1%). The city is divided into 10 boroughs, called sub-cities. Addis covers 527 square kilometres of area in Ethiopia. The population density is estimated to be near 5,165 individuals per square kilometre (18).

Addis Ababa is located at 9°1’48’’N 38°44’24’’E. It has an elevation of 2,326 meters above sea level, at the lowest point around Bole International Airport in the southern periphery and it rises to over 3,000 metres above sea level in the Entoto Mountains to the North. The large-scale assessment of the knowledge and practice in the prevention of COVID-19 was conducted from 14 to 30 April, 2020.

### Study population

A population assumed to be more likely within high risk groups of acquiring COVID-19 was the study population. More likely high-risk groups are classified as groups who have high exposure due to different risk factors of COVID-19 such as, exposure to public areas, travellers, recently been overseas, close contact of COVID-19 positive confirmed cases, and being in correctional and detention facilities. Health workers (including emergency services and non-clinical staff) regardless of their risk, and street children, prisons, schools, long-term living facilities, bus stations workers etc) were also classified as a high-risk.

### Study participants

Most of the respondents were formal sector employees. These formal sector target facilities included; the bus stations, public transport drivers, airport, cargo, health facilities, public, and private pharmacies, hotels, government and private banks, telecom centers, trade centers, children’s villages, custody centers, geriatric centers, prison, detention centers and selected slum areas where people live in crowded housing (namely; Lideta, Atobis Tera, Kirkos).

### Sampling strategy and data collection

Random sampling method was applied to identify the study participants. In total, 6,007 people were included.

Background and demographic information was collected using a questionnaire that is adapted from the surveillance system for COVID-19. Senior statisticians developed the data entry template using ODK. Internet-based cloud was used to transfer the data and computer tablets were used for data collection. Information on reported symptoms, travel history, any suspicious exposure to COVID-19 and attending gatherings/crowded areas was collected. Thirty-two teams, each of them with two members, were involved in the fieldwork. Sixty-four health professionals (Medical Doctors, Public Health Officers, Laboratories, Nurses, Environmental health professional and Midwives) were trained for three days. The training included a classroom lecture and practical exercise on the use of the data collection tool. During the face-to-face interview, the field teams used all the necessary PPE. They also practiced all the necessary precautions such as practicing hand hygiene, to avert the risk of acquiring the infection from the study participants.

## Data source

The analysis was done on the data that was collected for the large-scale COVID-19 screening in Addis Ababa. The collected data includes socio-demographic information (such as age, sex, religion, and occupation), income, knowledge about the transmission and prevention methods. Personal practices towards the prevention of COVD-19 and reported symptoms were also captured.

## Definition of variables

The knowledge on the transmission of COVID-19 was measured using four items. The items were; 1) during contacts (handshake and physical contact), 2) during coughing and sneezing, 3) during breathing, and 4) Eating and drinking contaminated foods. Good knowledge was classified when the scoring was ≥3 (75% and above) out of four items and below 3 indicated poor knowledge on the transmission methods of Covid-19.

Knowledge on the prevention of COVID19 was measured using four items. The items were awareness on, 1) Physical distancing, 2) Hand washing, 3) Use of mask, and 4) Isolation and quarantine. Good knowledge of prevention was classified as when the scoring was ≥3 (75% and above) out of four items and score below 3 indicated poor knowledge on prevention methods of Covid-19.

The score of the practice was measured based on four items. These four items were, 1) practice of hand hygiene, 2) use of mask, 3) maintaining social distance and 4) staying home.

A score ≥3 (75% and above) were classified as a good practice towards the precautionary measures of COVID-19 and a score of ≤2 (50% and below) indicated a poor practice.

### Data Quality Control

To maintain the quality of the data, two days training was provided for data collectors on the aim of the study and methods of data collection. A computer-based data collection was used to control entry errors. Field level suppression was done to control the data collection and data quality.

### Data management and analysis

We obtained the data from EPHI. The data was collected for the routine surveillance and screening activity among more priority high risk groups of Addis Ababa people. Data were directly transferred from the field staff tablet to EPHI server whenever the field team completed each questionnaire.

The items were first coded as “1” favouring the good outcome and “0” not favouring a good outcome. Then, all the items were added into one to represent the outcome. The new variable had a total score range from 0 to 4. A score of ≥3 (75%) would favour and ≤2 (50%) would disfavour the outcome.

A descriptive statistic was done to present the results in count, proportion and graphs. Multivariate Logistic regression was done to determine the factors associated with knowledge on transmission and prevention of COVID-19 and practice towards the prevention of COVID-19.

## Result

### Socio-demographic characteristic of the participants

A total of 6,007 participants were involved in the survey. Of these, the majority, 61% (3,634) were male; 41% (2,470) were between ages 25-34 years; and around 4% (236) were above 60 years old. The mean age of the participants was 33(SD ±11.73) years. Over half (50%) of the participants were non-formal employees such as street children, clients living in centres, in children’s villages, cardiac centres and cancer centres, prisons etc, while a small proportion were health care workers (14%) and public transport drivers (11%). Around, three-quarters (75%) of the participants were Orthodox Christian in religion and half of the study participants (50%) had a monthly income between 3,001 and 10,000 ETB **(Table 1)**.

**Table 1.** Socio-demographic characteristics of the study participants, Addis Ababa, Ethiopia, 2020 (n=6,007)

| Characteristics | Categories | Number(n) | Percent (%) |
| --- | --- | --- | --- |
| Sex | Male | 3634 | 60.5 |
|  | Female | 2373 | 39.5 |
| Age (Years) | less than 15 | 55 | 0.9 |
|  | 15-24 | 1220 | 20.3 |
|  | 25-34 | 2470 | 41.1 |
|  | 35-44 | 1245 | 20.7 |
|  | 45-59 | 781 | 13.0 |
|  | 60+ | 236 | 3.9 |
| Occupation | Health care worker | 867 | 14.4 |
|  | Working with Animals | 49 | 0.8 |
|  | Driver | 683 | 11.4 |
|  | Guard | 547 | 9.1 |
|  | Kiosk/grocery | 316 | 5.3 |
|  | Student | 134 | 2.2 |
|  | Others | 3411 | 56.8 |
| Religion | Orthodox | 4500 | 74.9 |
|  | Muslim | 673 | 11.2 |
|  | Protestant | 670 | 11.2 |
|  | Catholic | 94 | 1.6 |
|  | Other | 70 | 1.2 |
| Income in ETB | <=1000 | 781 | 13.0 |
|  | 1001-3000 | 1793 | 29.8 |
|  | 3001-10000 | 3019 | 50.3 |
|  | >10000 | 414 | 6.9 |

### Experience of COVID 19 related illness and communication

The experiences of COVID-19 related illness and communication were assessed. Among the study participants, 3% had a history of fever and an experience of respiratory illness within 14 days before the study. Of those who had an experience of respiratory illness, 51% had a cough, 34% had a runny nose, 18% had a sore throat and the rest, 11%, had shortness of breath. Free call centres and hotlines have been made publicly available to create awareness, receive information and for other COVID-19 related issues; however, only 11% of participants contacted these facilities.

### Knowledge of participants on the transmission, prevention, and practice for prevention of COVID19

The majority, 87%, of the respondents mentioned direct contact with COVID-19 patients, and 71% mentioned breathing as possible transmission mechanisms for COVID-19. Regarding prevention, 85% and 83%mentioned hand washing and social distancing as prevention mechanisms for COVID-19, respectively. More than 80% of the respondents practiced hand washing, while 76% practiced social distancing to prevent the transmission of COVID-19 (**Table 2**).

**Table 2.**
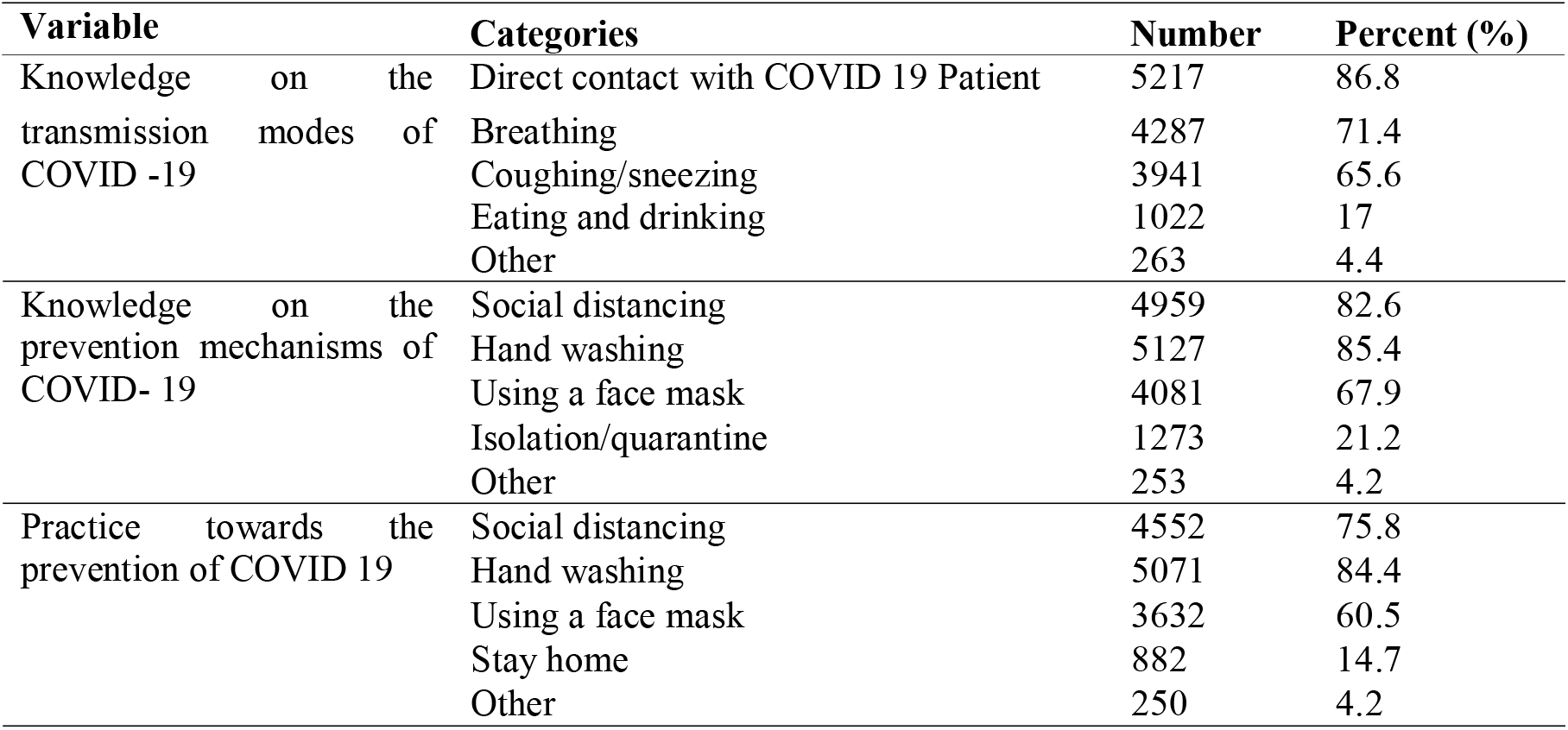
Knowledge on transmission, prevention and practice towards the prevention of COVID 19 among most likely to be high risk groups in Addis Ababa, Ethiopia, 2020

### Level of knowledge on transmission modes and prevention methods, and practice for the prevention of COVID-19

Findings demonstrated that study participants have low levels of knowledge on transmission modes, prevention methods and practice of prevention of COVID-19. Over half (52%; 95% CL: 51-54) of the respondents had good knowledge on transmission of COVID-19; about 60% (95% CL: 58-61) had good knowledge on prevention; and about half, 49% (95% CI: 48-50) had poor practices of the precautionary measures against COVID-19 **(Fig 1)**.

**Figure 1.**
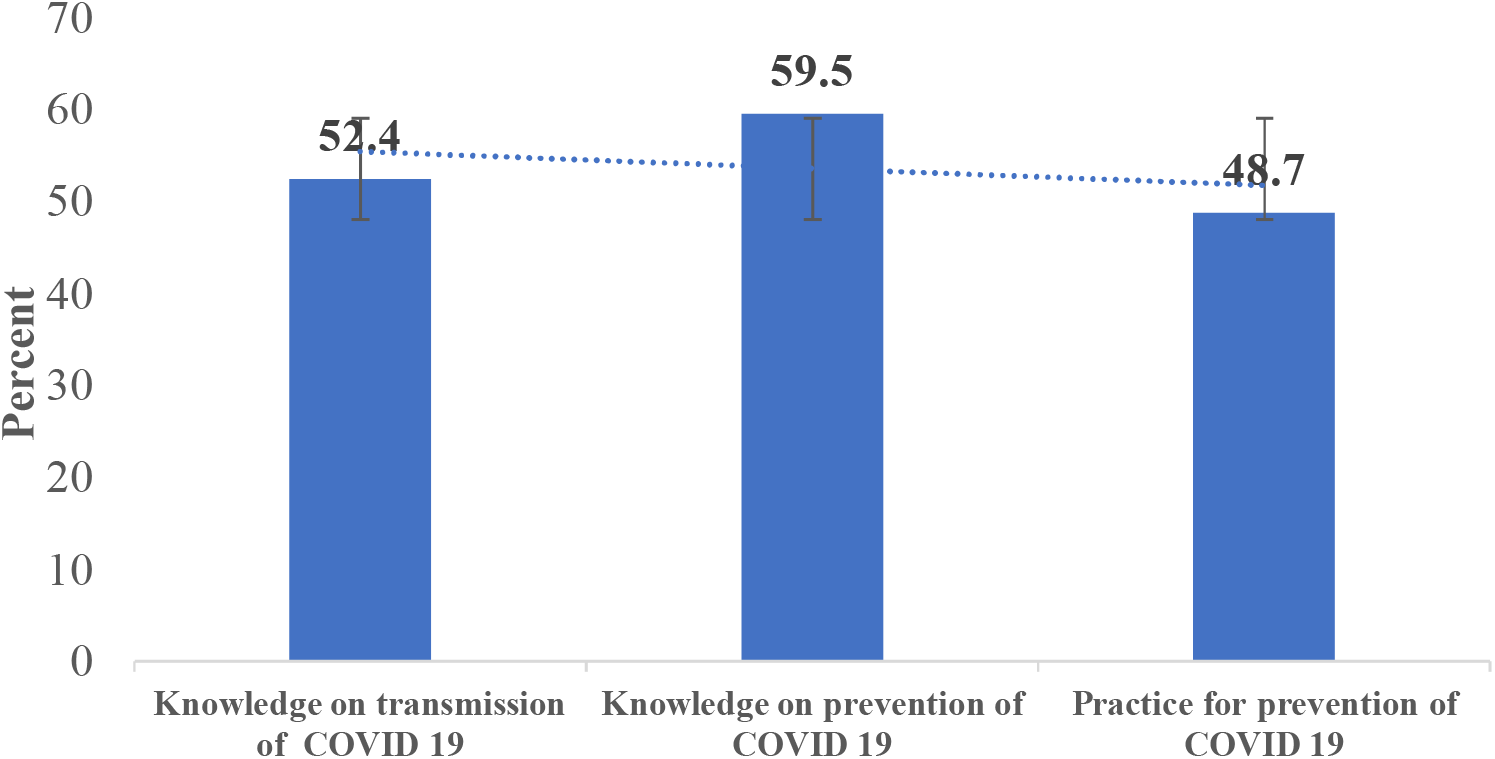
Level of knowledge on the transmission, prevention and practice of precautionary measure of COVID-19

### Factors associated with knowledge on the transmission mechanisms of COVID 19

Multiple logistic regression analysis showed that gender, age, occupation, income and having contacted the 8335 free call center were significantly associated with knowledge on the transmission mechanisms of COVID-19. The odds of being knowledgeable on the transmission mechanisms of COVID-19 among male participants were 0.87 times less likely than females (AOR= 0.87, 95% CI: 0.78 - 0.97). As age increases, the likelihood of being knowledgeable about the transmission modes of COVID-19 increases. Age group > 64 years were 14 times (AOR: 14.38, CI= 6.79 - 30.48) more likely to be knowledgeable on the transmission of COVID-19 than those who were younger than 18 years old. Regarding the occupation of respondents, health workers and grocery store workers were 2.25 and 2.22 times, respectively, more likely to be knowledgeable on the transmission methods of COVID-19 than other occupations. As income increases, the likelihood of being knowledgeable about the methods of transmission of COVID also increases; however, above some level of income this is no longer the case. Participants whose income was greater than 10,000 Ethiopian Birr were 49% (AOR: 0.51, 95%CI=0.39 - 0.65) less likely to be knowledgeable about the transmission mechanism of COVID-19 than low-income participants (<1000 ETB). Those participants who had contacted 8335 free call center were 1.3 times more likely to be knowledgeable about transmission mechanisms of COVID-19 than those who did not.(AOR=1.30, 95% CI=1.10 - 1.54) (**Table 3**).

**Table 3.**
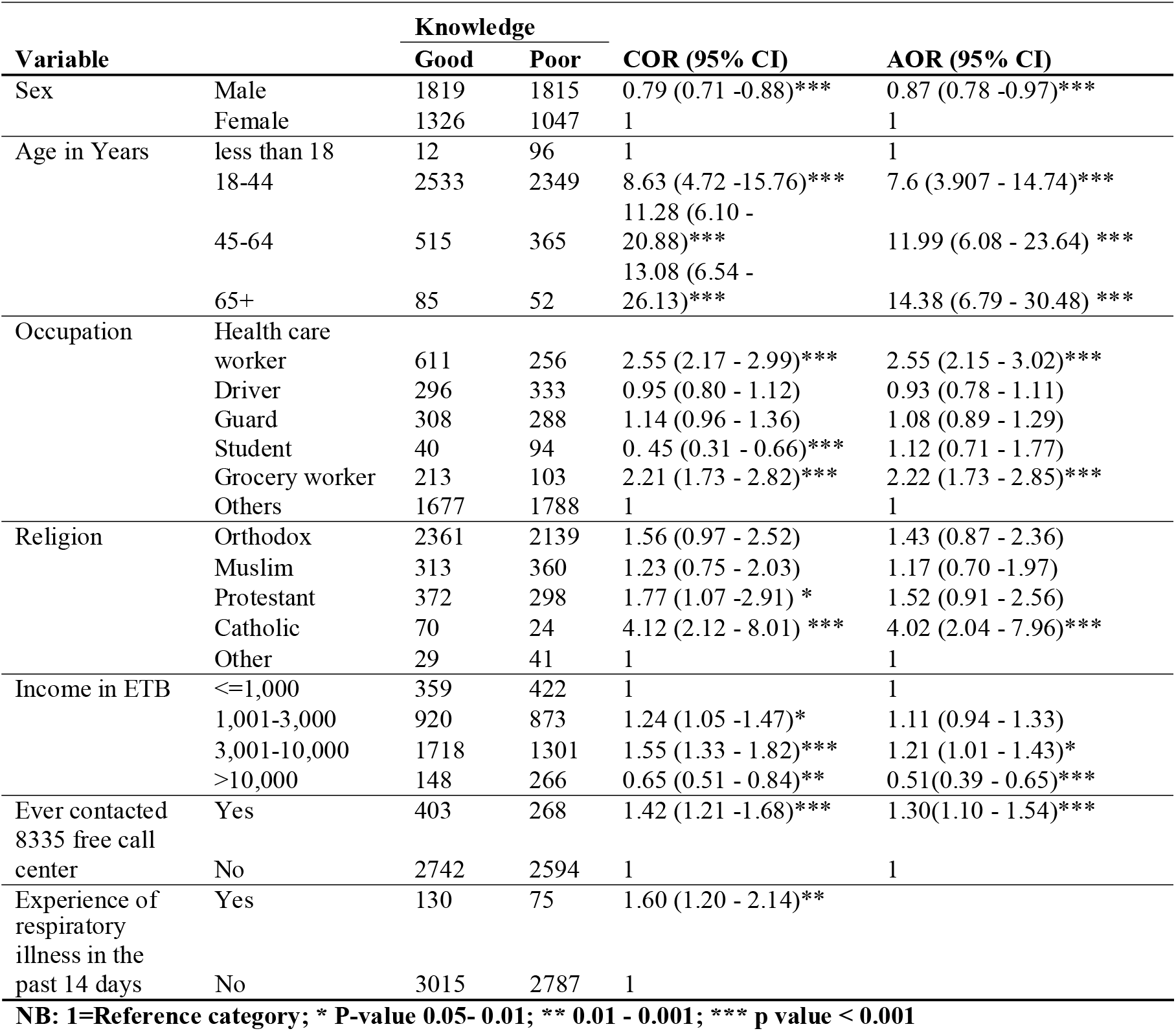
Factors associated with knowledge on the transmission of COVID 19 among participants, Addis Ababa, Ethiopia, 2020.

| Variable |  | Knowledge |  | COR (95% CI) | AOR (95% CI) |
| --- | --- | --- | --- | --- | --- |
|  |  | Good | Poor |  |  |
| Sex | Male | 1819 | 1815 | 0.79 (0.71 -0.88)*** | 0.87 (0.78 -0.97)*** |
|  | Female | 1326 | 1047 | 1 | 1 |
| Age in Years | less than 18 | 12 | 96 | 1 | 1 |
|  | 18-44 | 2533 | 2349 | 8.63 (4.72 -15.76)*** | 7.6 (3.907 - 14.74)*** |
|  | 45-64 | 515 | 365 | 11.28 (6.10 - 20.88)*** | 11.99 (6.08 - 23.64) *** |
|  | 65+ | 85 | 52 | 13.08 (6.54 - 26.13)*** | 14.38 (6.79 - 30.48) *** |
| Occupation | Health care worker | 611 | 256 | 2.55 (2.17 - 2.99)*** | 2.55 (2.15 - 3.02)*** |
|  | Driver | 296 | 333 | 0.95 (0.80 - 1.12) | 0.93 (0.78 - 1.11) |
|  | Guard | 308 | 288 | 1.14 (0.96 - 1.36) | 1.08 (0.89 - 1.29) |
|  | Student | 40 | 94 | 0.45 (0.31 - 0.66)*** | 1.12 (0.71 - 1.77) |
|  | Grocery worker | 213 | 103 | 2.21 (1.73 - 2.82)*** | 2.22 (1.73 - 2.85)*** |
|  | Others | 1677 | 1788 | 1 | 1 |
| Religion | Orthodox | 2361 | 2139 | 1.56 (0.97 - 2.52) | 1.43 (0.87 - 2.36) |
|  | Muslim | 313 | 360 | 1.23 (0.75 - 2.03) | 1.17 (0.70 -1.97) |
|  | Protestant | 372 | 298 | 1.77 (1.07 -2.91) * | 1.52 (0.91 - 2.56) |
|  | Catholic | 70 | 24 | 4.12 (2.12 - 8.01) *** | 4.02 (2.04 - 7.96)*** |
|  | Other | 29 | 41 | 1 | 1 |
| Income in ETB | <=1,000 | 359 | 422 | 1 | 1 |
|  | 1,001-3,000 | 920 | 873 | 1.24 (1.05 -1.47)* | 1.11 (0.94 - 1.33) |
|  | 3,001-10,000 | 1718 | 1301 | 1.55 (1.33 - 1.82)*** | 1.21 (1.01 - 1.43)* |
|  | >10,000 | 148 | 266 | 0.65 (0.51 - 0.84)** | 0.51(0.39 - 0.65)*** |
| Ever contacted 8335 free call center | Yes | 403 | 268 | 1.42 (1.21 -1.68)*** | 1.30(1.10 - 1.54)*** |
|  | No | 2742 | 2594 | 1 | 1 |
| Experience of | Yes | 130 | 75 | 1.60 (1.20 - 2.14)** |  |

|  |  |  |  |  |
| --- | --- | --- | --- | --- |
| respiratory<br>illness in the<br>past 14 days | No | 3015 | 2787 | 1 |

NB: 1=Reference category; \* P-value 0.05- 0.01; \*\* 0.01 - 0.001; \*\*\* p value < 0.001

### Factors associated with knowledge on the prevention of COVID-19

Multiple logistic regression analysis showed that age, occupation, income, religion, and experience of respiratory illness 14 days before the study were significantly associated with knowledge on the prevention of COVID-19. At age 64 or greater, knowledge on the prevention methods of COVID-19 was 11 times higher than at ages below 18 years old. (AOR=11.46, 95%CI=5.57 - 23.59). Regarding the occupation of respondents, health workers and grocery store workers were 4 and 2 times more likely, respectively, to be knowledgeable on the prevention methods, while drivers were 18%less likely to be knowledgeable about the prevention methods of COVID-19 than other occupants. Those participants who experienced respiratory illness in the past 14 days were 1.6 times (AOR=1.61, 95%CI =1.17 -2.21) more likely to be knowledgeable about the prevention methods of COVID-19 than those who did not experience respiratory illness **(Table 4)**.

**Table 4.** Factors associated with knowledge on the prevention of COVID 19 among participants, Addis Ababa, Ethiopia, 2020

| Variable |  | Knowledge |  | COR (95% CI) | AOR (95% CI) |
| --- | --- | --- | --- | --- | --- |
|  |  | Good | Poor |  |  |
| Age in Years | less than 18 | 14 | 94 | 1 | 1 |
|  | 18-44 | 2902 | 1980 | 9.84 (5.56 - 17.30) | 8.12(4.32 - 15.26)*** |
|  | 45-64 | 575 | 305 | 12.66 (7.10 - 22.57) | 12.42 (6.50 - 23.74)*** |
|  | 65+ | 85 | 52 | 10.98 (5.68 - 21.21) | 11.46 (5.57 - 23.59)*** |
| Occupation | HCWs | 723 | 144 | 4.14 (3.42 - 5.01)*** | 4.27 (3.5 - 5.21)*** |
|  | Drivers | 319 | 310 | 0.85 (0.72 - 1.01) | 0.82 (0.69 - 0.97)* |
|  | Guards | 358 | 238 | 1.24 (1.04 - 1.48) * | 1.14 (0.96 - 1.37) |
|  | Students | 46 | 88 | 0.43 (0.3 - 0.62) *** | 1.19 (0.76 - 1.87) |
|  | Grocery workers | 231 | 85 | 2.24 (1.73 - 2.89)*** | 2.32 (1.78 - 3.01)*** |
|  | Others | 1899 | 1566 | 1 | 1 |
| Religion | Orthodox | 2685 | 1815 | 1.66 (1.03 - 2.66)*** | 1.62 (0.98 - 2.67) |
|  | Muslim | 362 | 311 | 1.31 (0.79 - 2.14) | 1.29 (0.77 - 2.16) |
|  | Protestant | 424 | 246 | 1.93 (1.18 - 3.17)** | 1.68 (0.99 - 2.83) |
|  | Catholic | 72 | 22 | 3.67 (1.88 - 7.17)*** | 4.04 (2.02 - 8.06) *** |
|  | Other | 33 | 37 | 1 | 1 |
| Income in ETB | <=1000 | 376 | 405 | 1 | 1 |
|  | 1001-3000 | 1071 | 722 | 1.60 (1.35 - 1.89)*** | 1.42 (1.19 - 1.69)*** |
|  | 3001-10000 | 1912 | 1107 | 1.86 (1.59 - 2.18)*** | 1.31 (1.10 - 1.55)** |
|  | >10000 | 217 | 197 | 1.19 (0.94 - 1.51) | 0.86 (0.67 - 1.10) |
| Experience of respiratory illness in the past 14 days | Yes | 146 | 59 | 1.71 (1.26 - 2.33)** | 1.61 (1.17 - 2.21)** |
|  | No | 3430 | 2372 | 1 | 1 |

**NB: 1=Reference category; \* P Value 0.05- 0.01; \*\* 0.01-0.001; \*\*\* p value < 0.001**

Using logistic regression analysis, the study found that occupation, religion, income, and knowledge on the transmission and prevention of COVID-19 were found to be associated with the practice of precautionary measures against COVID-19.

As depicted in table 5, health care workers were more than two times (AOR=2.33 (95%IC: 1.85 - 2.93) more likely to implement the precautionary measures of COVID-19 than other occupants. Also, drivers (AOR= 1.94, 95% CI: 1.49 -2.52) and guards (AOR=1.83, 95% CI: 1.40 - 2.39) had higher odds of implementing the precautionary measures of COVID-19, respectively. Implementation of precautionary measures against COVID-19 was found to be lower among grocery store workers, at 0.71 (95% CI: 0.53 - 0.95) and students at 0.56, (95% CI: 0.32 - 1.00) compared to other occupants. Regarding religion, Muslims participants were at0.42 (95% CI: 0.19 - 0.88) lower odds of implementing the precautionary measures of COVID-19 than others. The implementation of precautionary measures against COVID-19 were found to be lower among higher income groups. Those who earned more than 10,000 ETB per month had 0.42 lower odds of practicing the precautionary measures against COVID-19 than those with lower income.

**Table 5.**
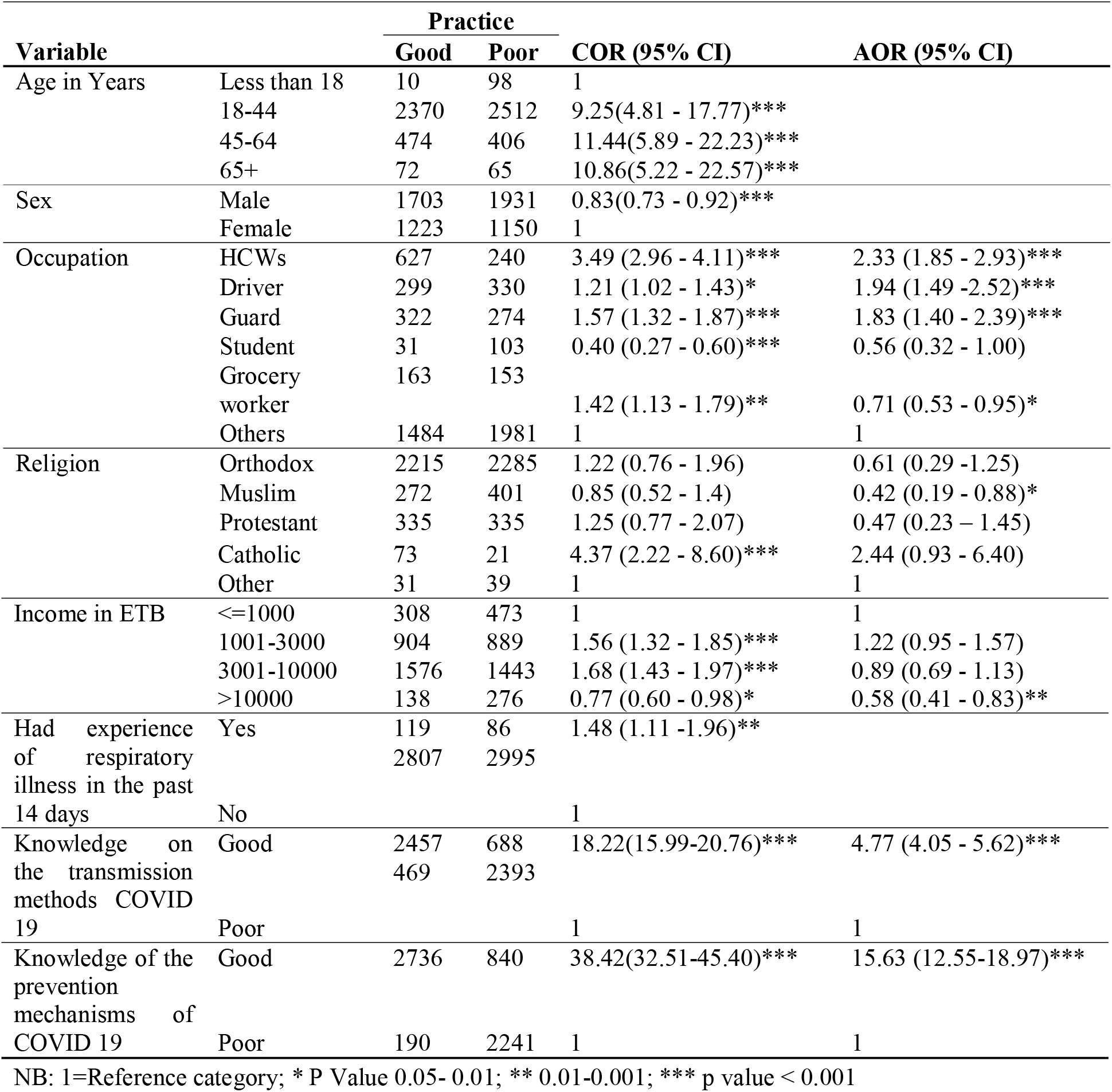
Factors associated with the practice of precautionary measures against COVID 19 among participants, Addis Ababa, Ethiopia, 2020

Participants who had good knowledge on transmission methods and prevention mechanisms COVID-19 were four times (95% CI: 4.05 - 5.62) and fifteen times (95% CI: 12.55 -18.97) more likely, respectively, to implement the precautionary measures of COVID-19**(Table 5**).

## Discussion

The study found 52% and 60% overall knowledge of the respondents about transmission and prevention of COVID-19, respectively. The practice of the prevention of COVID-19 was low (49%). Regarding the factors, gender, age, occupation, income and having used the 8335 free call center were the factors associated with knowledge on the transmission of COVID-19. Age, occupation, income, religion, and experience of respiratory illness 14 days before the study were significantly associated with the knowledge on the prevention of COVID-19. The study found that occupation, religion, income, and knowledge on the transmission and prevention of COVID-19 were associated with the practice of precautionary measures against COVID-19.

This study was conducted two months after the first COVID-19 positive case was reported in the country; thus we expect the study community to have higher awareness now. However, the level of knowledge that this study identified was unexpectedly low. The Ethiopian study is compared and contrasted to some others globally. A study conducted in Iran revealed that knowledge about the mode of transmission of the disease found a similar figure of 56% (8). In contrast, a study conducted in China revealed that the overall knowledge of COVID-19 was high at 90% (16). Our study has found better knowledge on the prevention of COVID-19 than a study conducted in Thailand with a result of, 73.4% on the knowledge on prevention and control of COVID-19 (19).

The level of practices for the prevention of COVID-19 in Ethiopia was 49%, which was lower than the study conducted in Iran, 71%, (8),but much higher than the study conducted in Thailand, 17%, (19). The sociodemographic characteristics of the study participants, study differences in methodology, study time, and area of the study could explain the differences, while the low-levels of use of the 8335 call centers and hotline may have limited people from acquiring knowledge in the subject matter.

In this study, age, occupation, and income have shown significant association with the knowledge on the transmission and prevention mechanisms of COVID-19. Gender, religion, and contacting the 8335 free call center were significantly associated with the knowledge on the transmission mechanisms of COVID-19, while participants who had a respiratory illness 14 days before the study were significantly associated with knowledge on the prevention mechanisms of COVID-19. The findings of women being more knowledgeable than men about the transmission of COVID-19 was in line with the studies conducted in China and Iran (8,16).

Being a health professional was associated with better knowledge about COVID-19. This finding is supported by the study conducted in Iran (8). A further similarity was found between two studies on the association between age groups and knowledge, with both studies showing better knowledge at older ages. The increase in income was inversely associated with being knowledgeable about the transmission methods of COVID-19. In addition, the study revealed that those participants who had contacted the 8335 free call center had better knowledge on the transmission methods of COVID-19, and those patients who have experience on respiratory illness 14 days before the study had better knowledge on the prevention of COVID-19.

The study also found that occupation, religion, income, and knowledge on the transmission and prevention of COVID-19 were found to be associated with the practice of precautionary measures against COVID-19. Health care workers, drivers, and guards reported practicing prevention mechanisms such as hand washing, social distancing, use of face masks, and staying home. On the other hand, participants such as grocery store workers, students and high-income groups were not practicing the prevention methods of COVID-19 in the proper ways. Contrary to our study, having health-care-related occupations was found to be significantly associated with lower practice towards the prevention of COVID-19 (8). The hand hygiene practice has strong relationship and it is widely practiced among Muslim religion followers. However, the measurement of precautionary measures was estimated in combined with other three preventive practices that lead to pulling the result towards negative; such as the use of mask, physical distancing and staying at home. Our study revealed that having good knowledge on the transmission and prevention of COVID-19 was found to be associated with good practices on COVID-19 prevention. This finding is not in line with the study conducted in China, which revealed that study participants who had a good knowledge score on the prevention mechanisms of COVID-19 were more likely to visit crowded places and failed to wear face masks (16).

### Strengths and limitations

Our study was large and had a broad scope with several strengths. It was based on a large sample size and focused on participants from areas that were likely to be high risk; therefore, the findings can be generalized across the city’s population. Unlike other studies, we did face-to-face interviews in this challenging situation, which is another strength. However, it has also limitations. The study did not gather important characteristics of the study participants, such as educational status, marital status and visiting public places, smoking etc that could affect knowledge on COVID19. Further, the initial assessment was done as part of the routine COVID-19 response rather than specifically for this study.

### Conclusion

The study pointed out that respondents had moderate knowledge about transmission modes, preventions mechanisms and practice on the prevention of COVID-19 areas of high risk in Addis Ababa. Awareness creation programs on more targeted groups, such as men, youth and those with higher incomes are required in order to improve the knowledge and practices on the prevention of COVID 19. Advocacy of the hand hygiene, physical distancing, use of mask and others strategies should fulfil the characteristics of the targeted individuals.

## Data Availability

Data can be made available at a reasonable request to the authors, Atkure Defar,
or Gebeyaw Molla

List of abbreviations
CI: Confidence Interval
COVID-19: Coronavirus disease 2019
ETB: Ethiopian Birr
EPHI: Ethiopian Public Health Institute
ODK: Open Data Kit
SD: Standard Deviation
PPE: Personal Protective Equipment

## Declaration

### Ethics approval and consent to participate

We obtained ethical approval from Institutional Review Board of the Ethiopian Public Health Institute (Ethics Ref: EPHI 6.13/789). During the data collection, written informed consent was obtained from each participants.

### Availability of data

Data can be made available at a reasonable request to the authors, Atkure Defar, or Gebeyaw Molla

### Authors’ contributions

AD, GM, AT and GT developed the idea of this analysis. GM analysed the data and AD, GM and AT drafted the paper. All authors contributed to the interpretation of results, revision of the manuscript, and approval of the final version of the paper.

### Consent for publication

Not applicable

## Acknowledgements

The authors would like to thank the EPHI for the support provided to the large-scale screening program. We also would like to thanks Albab Seifu and Hileena Eshetu Chole for proofreading the manuscript. Special thanks to all field staff for their excellent data collection and the study participants for their kind cooperation in interviews and data collection.

## Reference

1. World Health Organization (WHO). Coronavirus Disease (COVID-19) Outbreak: Rights, Roles And Responsibilities Of Health Workers, Including Key Considerations For Occupational Safety. 2019. p. 1–3. https://www.who.int/docs/default-source/coronaviruse/who-rights-roles-respon-hw-covid-19.pdf?sfvrsn=bcabd401_0

2. Qian X, Ren R, Wang Y, Guo Y, Fang J, Wu ZD, et al. Fighting against the common enemy of COVID-19: A practice of building a community with a shared future for mankind. Infect Dis Poverty. 2020;9(1):4–9.

3. World Health Organization 2020. Preparedness, prevention and control of COVID-19 in prisons and other places of detention [Internet]. Geneva; 2020. http://www.euro.who.int/data/assets/pdf_file/0019/434026/Preparedness-prevention-and-control-of-COVID-19-in-prisons.pdf

4. Wu Y, Chen C, Chan Y. The outbreak of COVID-19 J: An overview. J Chin Med Assoc. 2020;83:217–20.

5. Omer S, Ali S, Babar D. Preventive measures and management of COVID-19 in pregnancy. Drugs Ther Perspect [Internet]. 2020;(April):1–5. Available from: https://doi.org/10.1007/s40267-020-00725-x

6. Worldmeter report. COVID-19 CORONAVIRUS PANDEMIC. 2020. https://www.worldometers.info/coronavirus/?utm_campaign=CSauthorbio? Acceced date August 4/2020

7. Federal Mnistry of Health (FMOH) and Ethiopian Public Health Institute (EPHI). Notification Note on COVID-19 Situational Update. In: Notification Note. Addis Ababa, Ethiopia; 2020. https://www.ephi.gov.et/index.php/public-health-emergency/novel-corona-virus-update Acceced date June 4/2020.

8. Erfani A, Shahriarirad R, Ranjbar K, Mirahmadizadeh A, Moghadami M. Knowledge, Attitude and Practice toward the Novel Coronavirus (COVID-19) Outbreak: A Population-Based Survey in Iran. Bull World Heal Organ. 2020. https://www.who.int/bulletin/online_first/20-256651.pdf

9. Saqlain M, Munir MM, Rehman S-U, Gulzar A, Ahmed Z, Tahir AH, et al. Knowledge, Attitude and Practice among Healthcare Professionals regarding COVID-19: A cross-sectional survey from Pakistan. 2020. https://www.medrxiv.org/content/10.1101/2020.04.13.20063198v1.full.pdf

10. World Health Organization (WHO). Safe Ramadan practices in the context of the COVID-19. 2020;(April):1–3. https://apps.who.int/iris/bitstream/handle/10665/331767/WHO-2019-nCoV-Ramadan-2020.1-eng.pdf

11. FMOH. National Comprehensive Covid19 Management Handbook. First edit. Addis Ababa, Ethiopia; 2020. 1–102 p. Available from: http://www.moh.gov.et/ejcc/sites/default/files/2020-04/COVID_19 Handbook for health professionals FMOH 2020.pdf

12. Abuya AT, Austrian K, Isaac A, Kangwana B, Mbushi F, Muluve E, et al. COVID-19-related knowledge, attitudes, and practices in urban slums in Nairobi, Kenya. 2020;1–7. https://www.popcouncil.org/uploads/pdfs/2020PGY_Covid_KenyaKAPStudyDescription.pdf

13. Wolf MS, Serper M, Opsasnick L, Conor RMO, Curtis LM. Awareness, Attitudes, and Actions Related to COVID-19 Among Adults With Chronic Conditions at the Onset of the U.S. Outbreak. Ann Intern Med [Internet]. 2020;1–11. https://annals.org/aim/fullarticle/2764612/awareness-attitudes-actions-related-covid-19-among-adults-chronic-conditions

14. Iao H, Thi N, Han N, Khanh T Van, Ngan VK, Tam V Van, et al. Knowledge and attitude toward COVID-19 among healthcare workers at Knowledge and attitude toward COVID-19 among healthcare workers at District 2 Hospital, Ho Chi Minh City. Asian Pac J Trop Med. 2020;13(April):1–7.

15. Bhagavathula A, Bandari DK, Javad M, Mahabadi A, Bandari DK. Knowledge and Perceptions of COVID-19 Among Health Care Workers J: Cross-Sectional Study. JMIR PUBLIC Heal Surveill. 2020;6(2):1–9.

16. Zhong B, Luo W, Li H, Zhang Q, Liu X, Li W, et al. Knowledge, attitudes, and practices towards COVID-19 among Chinese residents during the rapid rise period of the COVID-19 outbreak: a quick online cross-sectional survey. Int J Biol Sci. 2020;16(10):1745–52.

17. Wadood A, Mamun A, Rafi A, Islam K, Mohd S, Lee LL, et al. Knowledge, attitude, practice and perception regarding COVID-19 among students in 2 Bangladesh: Survey in Rajshahi University. 2020. https://www.medrxiv.org/content/10.1101/2020.04.21.20074757v1.full.pdf+html. Acceced date May 31/2020

18. UN World Urbanization Prospects. Addis Ababa Population 2020. Addis Ababa; 2020. Available from: https://worldpopulationreview.com/world-cities/addis-ababa-population/%0AAddis

19. Srichan P, Apidechkul T, Tamornpark R, Yeemard F, Khunthason S, Kitchanapaiboon S, et al. Knowledge, attitude and preparedness to respond to the 2019 novel coronavirus (COVID-19) among the bordered population of northern Thailand in the early period of the outbreak: a cross-sectional study. 2020. https://papers.ssrn.com/sol3/papers.cfm?abstract_id=3546046

